# They’re Dying in the Suburbs: COVID-19 Cases and Deaths by Geography in Louisiana (USA)

**DOI:** 10.1101/2020.10.28.20221341

**Authors:** Alina Schnake-Mahl, Usama Bilal

## Abstract

The national COVID-19 conversation in the US has mostly focused on urban areas, without sufficient examination of another geography with large vulnerable populations: the suburbs. While suburbs are often thought of as areas of uniform affluence and racial homogeneity, over the past 20 years, poverty and diversity have increased substantially in the suburbs. In this study, we compare geographic and temporal trends in COVID-19 cases and deaths in Louisiana, one of the few states with high rates of COVID-19 during both the spring and summer. We find that incidence and mortality rates were initially highest in New Orleans. By the second peak, trends reversed: suburban areas experienced higher rates than New Orleans and similar rates to other urban and rural areas. We also find that increased social vulnerability was associated with increased positivity and incidence during the first peak. During the second peak, these associations reversed in New Orleans while persisting in other urban, suburban, and rural areas. The work draws attention to the high rates of COVID-19 cases and deaths in suburban areas and the importance of metropolitan-wide actions to address COVID-19.

**Registration:** N/A

**Funding source:** NIH (DP5OD26429) and RWJF (77644)

**Code and data availability:** Code for replication along with data is available here: https://github.com/alinasmahl1/COVID_Louisiana_Suburban/.

## INTRODUCTION

During the first presidential debate on September 29th, 2020, Vice President Biden called attention to the threat of COVID-19 in a location often overlooked in conversations about the epidemic: “they’re dying in the suburbs.” Evidence suggests the Vice President was correct (McMinn et al., 2020); in the spring and early summer, suburban areas experienced similar infection and deaths rates compared to urban areas (Zhang and Schwartz, 2020), but coverage has mostly focused on cities and urban areas as COVID-19 hot spots (Oster et al., 2020). In April, rates were highest in cities such as New York City, New Orleans (NOLA), and Seattle, but subsequent spread has moved beyond cities, and even in April, city boundaries did not cleanly contain infections (Frey, 2020; Oster et al., 2020).

This study examines geographic and temporal trends in COVID-19 cases and deaths in Louisiana, the state with some of the highest cumulative case rates per 100,000 people and one of few states that has already experienced two COVID-19 peaks (by October 2020). We first explore differential trends in positivity, incidence, and mortality between NOLA, other urban, suburban, and rural areas during the first peak, re-opening phase, and second peak in Louisiana. Second, we describe changing trends in the association between neighborhood social vulnerability and positivity and incidence, by geographic area.

## STUDY DATA AND METHODS

We used weekly census-tract (proxy for neighborhood) tests, positive tests, and confirmed case data made publicly available by the Louisiana Department of Health (LDH), and reported from February 27th to September 30^th^ (Louisiana Department of Health, 2020). We used daily parish-level (equivalent to county) mortality data, from the same period, from the Center for Systems Science and Engineering (CSSE) at Johns Hopkins University (Dong et al., 2020) because longitudinal mortality was not available from the LDH. We linked the census tract and parish data to the USDA 2010 Rural-Urban Commuting Area (RUCA) codes and Rural-Urban Continuum Codes (RUCC), respectively (USDA 2013, 2020). RUCA codes classify census tracts, and RUCC classify counties, both based on indicators of population density, urbanization, and adjacency to a metro area (for RUCC). We regrouped the codes into four geographies: New Orleans (NOLA) proper (Orleans parish/city), other urban, suburban, and rural areas (see **Appendix** for details). We designated NOLA separately because it is the most populous city in the state and has maintained differentially strict re-opening policies compared to other parishes (City of New Orleans, 2020). We also regrouped weeks into three periods: (1) first peak, encompassing March and April; (2) re-opening, starting in May (when LA initiated Phase 1 of re-opening) and going through June; and (3) second peak, starting in July, following the state’s movement into Phase 2 (**See Appendix Exhibit 4**).

We linked the census tract data to 2014-2018 American Community Survey population data to allow for rate calculations, and to the Centers for Disease Control Social Vulnerability Index (SVI), a summary index measuring populations most at risk during public health emergencies (Flanagan et al., 2011). We calculated the following monthly outcomes: incidence (confirmed cases/census tract population), positivity ratios (positive tests/total tests), and death rates (total deaths/parish population). To explore the association between SVI and census-tract outcomes, we calculated incidence and positivity by quintile of SVI. A higher SVI represents greater social vulnerability. We conducted all analyses in R version 4.0.2. We provide more detail on the data sources and analyses in the supplementary appendix.

## RESULTS

### Sample characteristics

We include data from 64 parishes and 1,105 census tracts. We designated 15% (174) of all census tracts as part of NOLA, 54% (591) as other urban, 23% (258) as suburban, and 10% (116) as rural; for parishes, the corresponding percentages were, 1.5%, 39%, 45%, and 14% respectively. Between February 27^th^ and September 30^th^, there were 2,026,674 total tests, 204,053 positive tests, 141,609 confirmed cases, and 5,321 deaths in Louisiana. For mortality, 587 deaths occurred in NOLA, 2,899 in other urban, 1,722 in suburban, and 113 in Rural, for respective rates of 15.1, 11.0, 11.2, and 10.9 per 10,000.

### Cases by geography

The proportion of NOLA cases decreased from the first (March to May 31^st^) to the second (June 1^st^-Sept 30th) half of the pandemic, while the percentage of cases in suburbs increased, and the proportion of rural and other urban cases remained stable (Exhibit 1). NOLA accounted for 14% of cases in the first half and 5% in the second half, while the suburban percentage increased from 18% to 26%.

**Exhibit 1:**
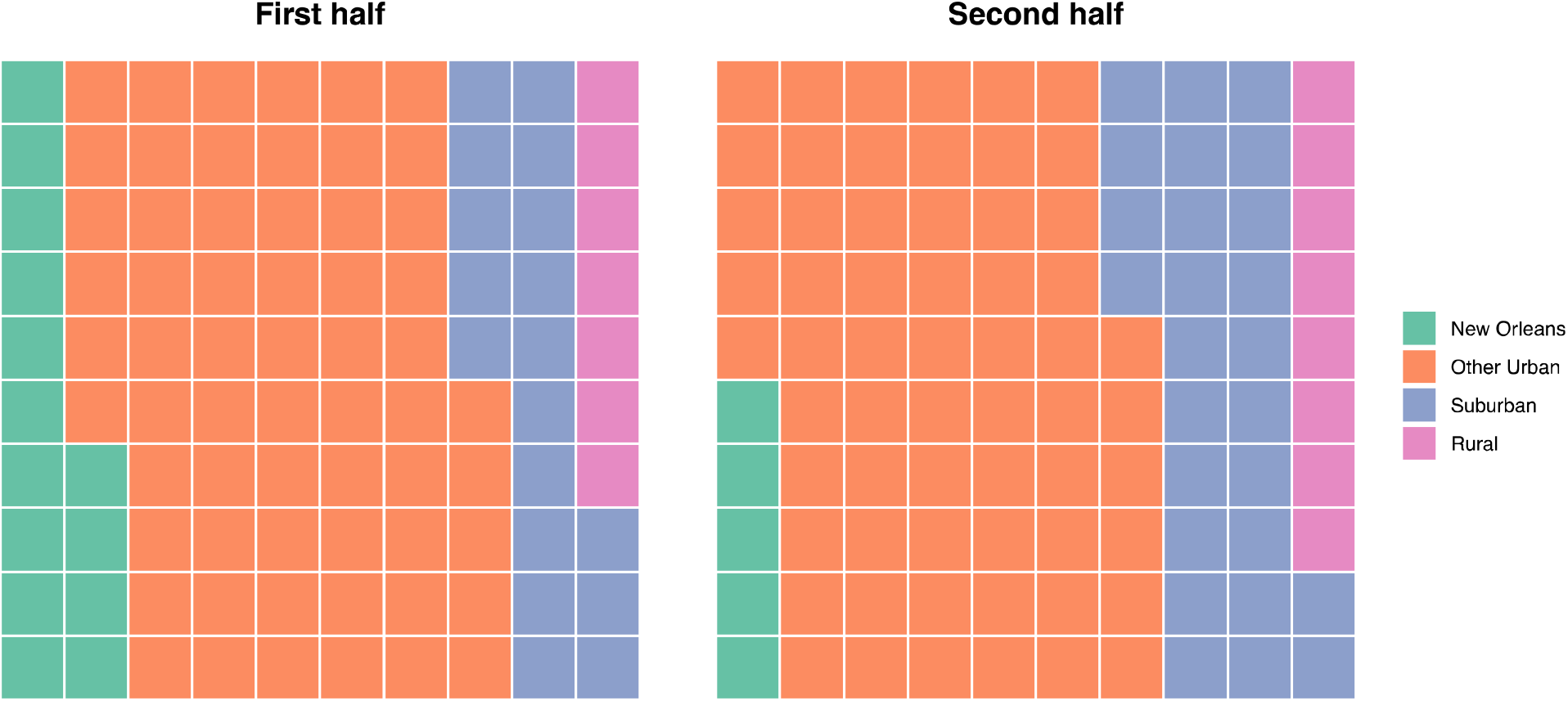
Percent of Cases by Geography in the First (March to May 31st) and Second (June 1st-Sept 30th) halfs of the Pandemic

### Trends by geography

Exhibit 2 shows temporal trends in positivity ratios, incidence rates, and mortality rates by geography. During the first peak, NOLA had the highest incidence (73 cases/10,000), positivity ratios (33%), and mortality rates (5.6 deaths/10,000). Other urban areas had 27 cases/10,000, followed by 16 and 11 cases per 10,000 in suburban and rural areas. Positivity ratios followed a similar pattern, at 20%, 18%, and 14% in other urban, suburban, and rural areas. Mortality rates in suburban and rural areas increased from 1.0 and 0.67 in the first peak to 2.4 and 2.2 per ten thousand in the second, while mortality rates in NOLA dropped substantially (7.3 to 0.5 per 10,000) and other urban rates dropped slightly (from 2.1 to 1.4 per 10,000). After the first peak, incidence rates declined for all geographies except rural areas, where incidence has increased steadily since the fall. In the second peak, incidence rates were over 70 cases/10,000 in suburban and other urban areas, nearly double the NOLA rate. Since May, positivity ratios have hovered close to 10% in other urban, suburban, and rural areas while remaining below 5% in NOLA.

**Exhibit 2:**
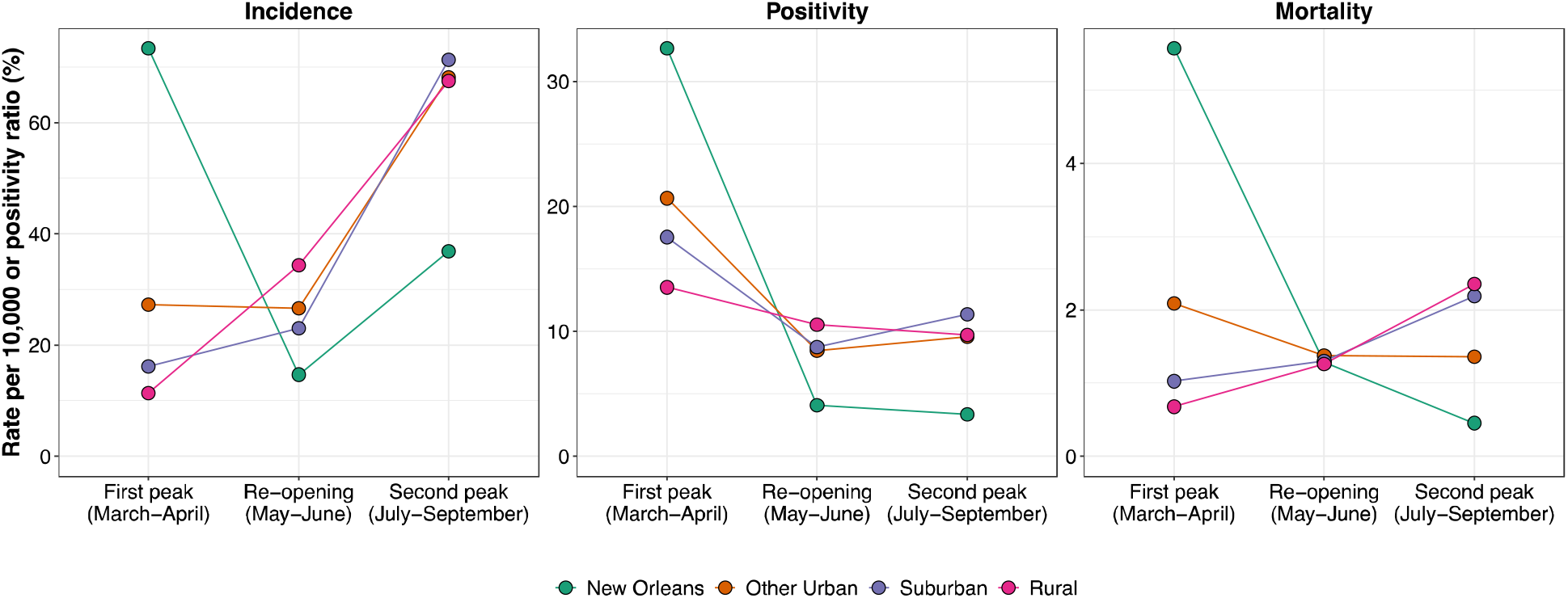
COVID-19 Outcomes by Geography in Louisiana.

### Social vulnerability and geography across peaks

During the first peak (March and April), positivity and incidence were higher in high social vulnerability neighborhoods. In NOLA, more than one in three (37%) tests were positive in neighborhoods in the highest social vulnerability quintile, as compared to less than one in four (22%) in neighborhoods in the lowest social vulnerability quintile. We did not observe a social gradient for either outcome in NOLA or rural areas during the reopening, and for other urban and suburban areas we found slightly higher positivity ratios and incidence rates in the most socially vulnerable areas. From July onwards, the second peak, shows no social gradient for any of the geographies for positivity, but an inverted social gradient for incidence in NOLA, with higher incidence in areas with lower social vulnerability. In other urban, suburban, and rural areas, we find a positive social gradient, where the most socially vulnerable areas show the highest incidence rates.

**Exhibit 3:**
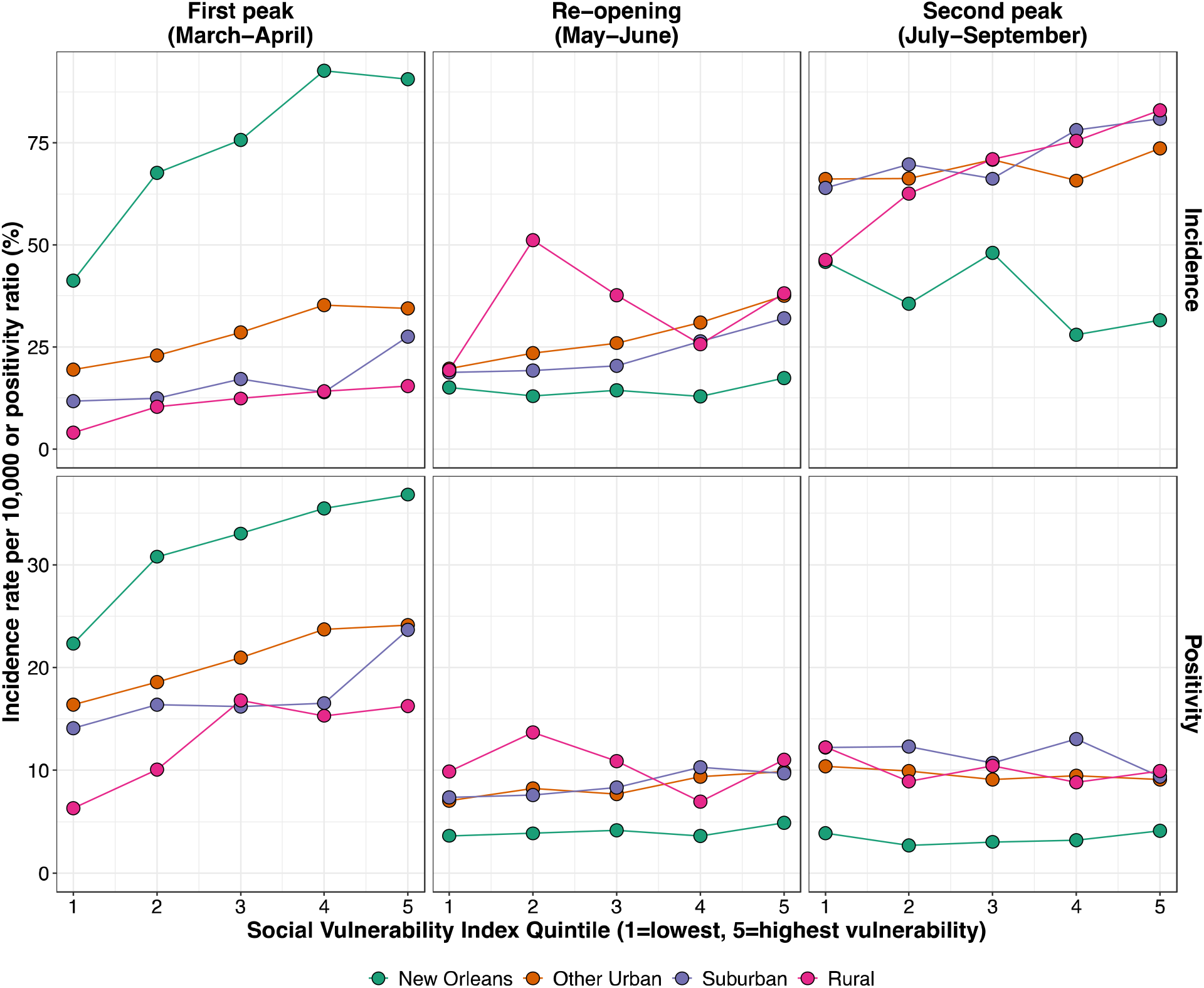
COVID-19 Outcomes by Geography and Social Vulnerability in Louisiana.

## CONCLUSIONS

Using census tract- and parish-level data in Louisiana, we found that following the first COVID-19 peak in March and April, incidence rates, positivity ratios, and mortality were higher in suburban, other urban, and rural areas as compared to NOLA, and these differences increased during the second peak. We also found disparities in incidence and positivity by social vulnerability during the first peak. These disparities were ameliorated during re-opening and the second peak, especially in NOLA, while they persisted in other urban, suburban, and rural areas. Importantly, positivity ratios were consistently highest in suburban areas through the summer and early fall, indicating either higher incidence or lower testing in suburban areas, especially as compared to NOLA. Finally, in the suburbs deaths rates for COVID-19 were over 10 per 10,000 residents, second only to NOLA. Together, this implies no protective effect of living in suburbs or rural areas on COVID-19 infection or overall mortality rates.

The high death rate in NOLA was likely driven by large case rates in the early months of the pandemic, during which case fatality rates were higher than in subsequent peaks (Fan et al., 2020). The lower incidence rates and positivity ratios in NOLA in the second peak may have resulted from more strict mitigation and suppression policies in the city (City of New Orleans, 2020), as changes in rates followed changes to policies (Yamana et al., 2020). Our findings also show lower incidence rates in more vulnerable neighborhoods of NOLA, while other highly vulnerable neighborhoods of Louisiana had higher incidence during the second peak. This suggests that stricter policies may protect the most vulnerable individuals, but our study cannot answer this causal question. Recent research has shown that racial disparities in COVID-19 mortality are mostly driven by differences in infection rates (Rentsch et al., 2020; Zelner et al., 2020). If this is the case, factors driving disparities in infection rates, such as occupational differences (McClure et al., 2020; Pirtle, 2020), may be positively impacted by mitigation policies. This evidence is consistent with other research, mostly in chronic diseases, showing that population-level interventions tend to reduce inequalities (Benach et al.; Frohlich, 2014; McLaren et al., 2010).

Health inequities result from a complex interplay of structural and systemic factors, including structural racism (Bailey et al., 2017; Williams, 2012) and economic inequality (Bor et al., 2017; Kawachi and Subramanian, 2014). These factors increase risk of SARS-COV-2 exposure and elevate the risk of severe COVID-19, and pattern the population distribution of COVID-19 outcomes (Bailey and Moon, 2020; Egede and Walker, 2020; Raifman and Raifman, 2020). Racial segregation, one of the primary mechanisms through which racism and economic inequality operate, separates populations by race/ethnicity and socioeconomic status, creating inequitable neighborhoods within metro areas (Williams and Collins, 2001). For most of the 20^th^ century, racial segregation allowed suburban areas to exist as largely white and middle class (Rothstein, 2017) through racist housing policies (Frey, 2011). However, over the last twenty years, suburbs’ demographics have shifted, increasing poverty and racial diversity in the suburbs (Allard, 2017; Kneebone and Berube, 2013). These recent sociodemographic changes suggest increasing vulnerability to chronic and infectious disease in the suburbs, given the strong relationship between social and economic marginalization and poor health outcomes (Chokshi, 2018). In addition, the early reopening and lack of adherence to social distancing and mask-wearing, which are common in more politically conservative areas (Katz et al., 2020; Rosenstrom et al., 2020), may have helped this increase in rates and mortality.

Patterns of high infection and death rates in Louisiana suburbs are likely generalizable to other states, especially to states beginning to experience a second infection peak. While studies have pointed to higher infection and spread of COVID-19 in metro compared to non-metro areas (Hamidi et al., 2020), and in more deprived neighborhoods of Louisiana (Oral et al., 2020), there has been limited examination of spatial inequities in COVID-19 between geographies. Given the economic and social interdependence of suburbs and cities, metropolitan-level COVID-19 policies can help reduce variability in outcomes and ensure access to testing and treatment across suburbs and cities, including 40% of the uninsured population who live in the suburbs (Schnake-Mahl and Sommers, 2017). The NOLA success in the second peak and high infection and positivity rates in nearby areas emphasize the importance of allowing local authority to determine stricter policies to protect their populations. As COVID-19 incidence increases again, suburban municipalities should consider imposing more stringent control policies in collaboration with nearby cities if the state declines to do so. Further research should consider how state preemption laws, a policy tool whereby a higher level of government prohibits or limits a lower government’s power to enact legislation (Haddow et al., 2020), impact overall COVID-19 rates and inequities between and within states and geographies.

Our analysis has several limitations. Like all US state data, the Louisiana COVID-19 data has a number of discrepancies, including backlogs to case reporting, that may impact the accuracy of monthly counts. The data only include tested cases, likely undercounting true cases, particularly early in the pandemic when testing access was limited. If access to testing differs by geography, for example, fewer testing sites in low-income suburban areas, the data may differentially undercount tests by geography. However, the positivity ratio should account for these differences.

## Conclusion

Our analysis provides a case study of spatial and temporal disparities in COVID-19, focused on the suburbs, a geography often overlooked regarding inequities. These results help call attention to areas impacted by the disease that may receive less media and academic attention, thereby helping policy makers target resources to areas in need and consider the policies most effective for the specifics of vulnerable populations, whether in suburban, urban, or rural areas. Future research should extend these geographic analyses to other states currently experiencing their second peaks.

## Data Availability

Code for replication along with data is available here: https://github.com/alinasmahl1/COVID_Louisiana_Suburban/

## APPENDIX

Data was assigned to census tract or parish, based on the resident of the individual tested, though census tracts with less than 1,000 people were not assigned cases to protect privacy; 140,357 of 168,512 (83%) tests could be matched to tracts. Death counts were not included for parishes with less than 25 deaths or that were not assigned to a parish, so our analysis excludes 190 deaths (3.4% of the total of 5 deaths).

**Appendix Exhibit 1: Primary and Alternative Geography Definitions**

### Definition

- Census Tracts (RUCA): New Orleans as all census tracts in Orleans Parish. Other Urban as metropolitan area core,, other than census tracts in Orleans Parish. Suburban includes metropolitan areas with high a low commuting, and micropolitan area core, high commuting, and low commuting. Rural areas are those not within a Metropolitan or Micropolitan area. See Appendix Exhibit 2.
- Parish (RUCC): New Orleans as New Orleans Parish. Other Urban as counties in metros with 250,000 or more. Suburban as metro counties of less than 250,000 and nonmetro counties with urban populations of more than 2,500. Rural as urban population of 2,500 to 19,999 not adjacent to a metro area, or completely rural, or less than 2,500 urban population. See Appendix Exhibit 3.

**Appendix Exhibit 2:** Rural Urban Commuting Area Codes (RUCA) Codes to Geographic Definitions.

| Code | Classification Description | Primary Definition |
| --- | --- | --- |
| 1 | Metropolitan area core: primary flow within an urbanized area (UA) | Other Urban (Other than NOLA) |
| 2 | Metropolitan area high commuting: primary flow 30% or more to a UA | Suburban |
| 3 | Metropolitan area low commuting: primary flow 10% to 30% to a UA | Suburban |
| 4 | Micropolitan area core: primary flow within an urban cluster of 10,000 to 49,999 (large UC) | Suburban |
| 5 | Micropolitan high commuting: primary flow 30% or more to a large UC | Suburban |
| 6 | Micropolitan low commuting: primary flow 10% to 30% to a large UC | Suburban |
| 7 | Small town core: primary flow within an urban cluster of 2,500 to 9,999 (small UC) | Rural |
| 8 | Small town high commuting: primary flow 30% or more to a small UC | Rural |
| 9 | Small town low commuting: primary flow 10% to 30% to a small UC | Rural |
| 10 | Rural areas: primary flow to a tract outside a UA or UC | Rural |
| 99 | Not coded: Census tract has zero population and no rural-urban identifier information | N/A |

**Appendix Exhibit 3:**
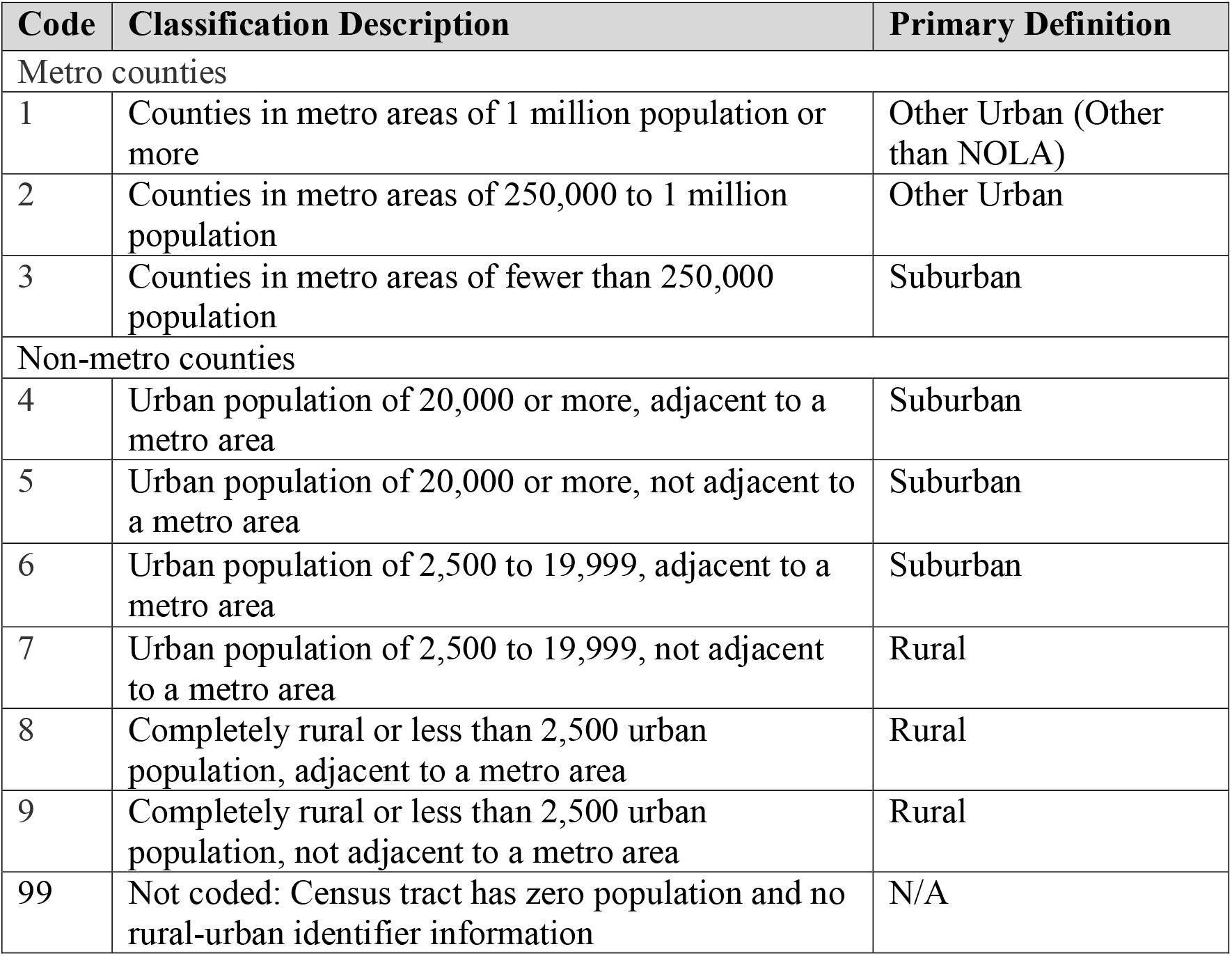
Rural Urban Continuum Codes (RUCC) to Geographic Definitions.

| Code | Classification Description | Primary Definition |
| --- | --- | --- |
| Metro counties |  |  |
| 1 | Counties in metro areas of 1 million population or more | Other Urban (Other than NOLA) |
| 2 | Counties in metro areas of 250,000 to 1 million population | Other Urban |
| 3 | Counties in metro areas of fewer than 250,000 population | Suburban |
| Non-metro counties |  |  |
| 4 | Urban population of 20,000 or more, adjacent to a metro area | Suburban |
| 5 | Urban population of 20,000 or more, not adjacent to a metro area | Suburban |
| 6 | Urban population of 2,500 to 19,999, adjacent to a metro area | Suburban |
| 7 | Urban population of 2,500 to 19,999, not adjacent to a metro area | Rural |
| 8 | Completely rural or less than 2,500 urban population, adjacent to a metro area | Rural |
| 9 | Completely rural or less than 2,500 urban population, not adjacent to a metro area | Rural |
| 99 | Not coded: Census tract has zero population and no rural-urban identifier information | N/A |

**Appendix Exhibit 4:**
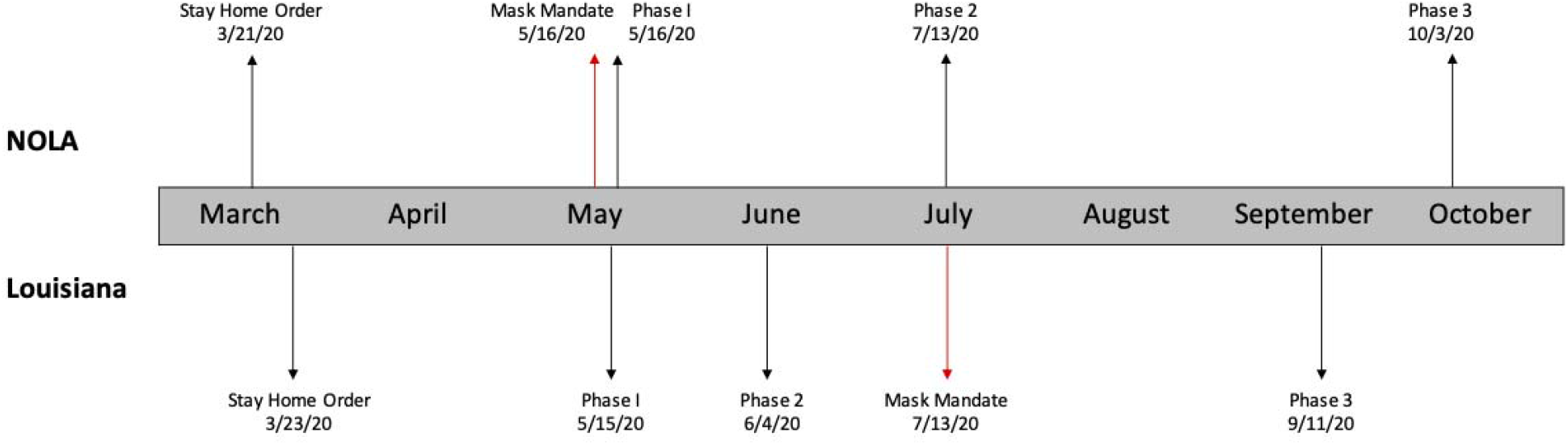
Louisiana and New Orleans COVID-19 Policy Timeline. \***Phase 1**: eased stay at home order and some restrictions; implemented occupancy limits, social distancing, and PPE requirements. **Phase 2:** allowed limited reopening for specific businesses, with capacity limitations and mask mandate (statewide) **Phase 3:** increased capacity for low-risk activities and further reopening of some non-essential businesses. New Orleans Phase 3 followed all of Louisiana’s guidelines and included additional local restrictions.

